# clickBrick Prompt Engineering: Optimizing Large Language Model Performance in Clinical Psychiatry

**DOI:** 10.1101/2025.06.28.25330267

**Authors:** Falk Gerrik Verhees, Fabian Huth, Vincent Meyer, Fabian Wolf, Michael Bauer, Andrea Pfennig, Philipp Ritter, Jakob Nikolas Kather, Isabella Catharina Wiest, Pavol Mikolas

**Author notes:** Corresponding author: Falk Gerrik Verhees, Department of Psychiatry and Psychotherapy Fetscherstraße 74, 01307 Dresden Germany. shared last authors (Isabella Catharina Wiest, Pavol Mikolas).

## Abstract

**Background:** Prompt engineering has the potential to enhance large language models’ (LLM) ability to solve tasks through improved in-context learning. In clinical research, the use of LLMs has shown expert-level performance for a variety of tasks ranging from pathology slide classification to identifying suicidality. We introduce clickBrick, a modular prompt-engineering framework, and rigorously test its effectiveness.

**Methods:** Here, we explore the effects of increasingly structuring prompts with the clickBrick framework for a comprehensive psychopathological assessment of 100 index patients from psychiatric electronic health records. We compare the performance of a locally-run LLM (Llama-3.1-70B-Instruct) against an expert-labelled ground truth for a variety of successively built-up prompts for the extraction of 12 transdiagnostic psychopathological criteria. Potential clinical value was explored by training linear support vector machines on outputs from the strongest and weakest prompts to predict discharge ICD-10 main diagnoses for a historical sample of 1,692 patients.

**Outcomes:** We could reliably extract information across 12 distinct psychopathological classification tasks from unstructured clinical text with balanced accuracies spanning 71 % to 94%. Across tasks, we observed a substantially improved extraction accuracy (between +19% and +36%) using clickBrick. The comparison unveiled great variations between prompts with a reasoning prompt performing best in 7 out of 12 domains. Clinical value and internal validity were approximated by downstream classification of eventual psychiatric diagnoses for 1,692 patients. Here, clickBrick led to an improvement in overall classification accuracy from 71% to 76%.

**Interpretation:** ClickBrick prompt engineering, i.e. iterative, expert-led design and testing, is critical for unlocking LLMs’ clinical potential. The framework offers a reproducible pathway for deploying trustworthy generative AI across mental health and other clinical fields.

**Funding:** The German Ministry of Research, Technology and Space and the German Research Foundation.

**Research in context:** *Evidence before this study:* We searched PubMed/MEDLINE articles without language restrictions published before June 25 2025 that combined three concept blocks - “prompt engineering” or related synonyms, “large language model/LLM” or specific model names (e.g., ChatGPT, GPT-4, LLaMA), and psychiatric or mental-health terms (e.g., psychiatry, psychotherapy, depression, anxiety). Additionally, we asked ChatGPT o3 to design and execute a systematic review strategy to also capture not-yet peer-reviewed but relevant pre-prints, given only our manuscript title. After manual de-duplication and abstract screening, three out of 23 identified studies did offer at least some information on their prompting strategies and were conducted on real-world clinical data from psychotherapy transcripts (one study on multi-dimensional counselling therapy, no peer review), or on online patient portal queries (two peer-reviewed studies on (a) empathy evaluation, and (b) provider satisfaction and use of generated response with partial integration with electronic health records. Neither systematically structured their prompts in a transparent way, nor tested reasoning prompts. Beyond psychiatry, one study analyzing automated echocardiography reports did employ a comparison between two different prompts and an expert-led design strategy. A single study used structured and transparent prompt engineering to generate automated responses for simulated problem-solving therapy sessions. None of the highlighted studies reported both head-to-head comparisons of competing prompt strategies for full reproducibility, and their application in real-world care, e.g. on electronic health records. Collectively, the existing literature suggests growing interest but reveals a paucity of rigorous evidence on how prompt engineering impacts large language model performance in clinical psychiatry, particularly in real-world settings.

*Added value of this study:* We demonstrate reliable information extraction from electronic health records across 12 distinct psychopathological classification tasks from unstructured clinical text and substantially improved extraction accuracy (between +19% and +36%) using clickBrick, our prompt engineering framework. The rationale for such an approach is justified by the surprising identification of zero-shot, few-shot and reasoning prompts as the best performing prompts for different tasks, while a Chain-of-Thought reasoning prompt performs best in 7 out of 12 tasks. And while most studies rely on proprietary language models like openAI’s ChatGPT, our locally run version of a popular open-weight model (Llama-3.1-70B-Instruct) allows for privacy safeguarding of sensitive patient data, which is essential for ethical clinical application.

*Implications of all the available evidence:* Generative artificial intelligence is poised to benefit psychiatric patients greatly, powering advances from therapy delivery to decision support and patient outreach. Rigorous prompt engineering with tools like clickBrick heightens their reliability and credibility, making clickBrick a cornerstone for bringing AI into everyday psychiatric care.

## Introduction

Generative artificial intelligence (AI) and large language models (LLMs) offer manifold applications across all domains of medicine from cancer classification in pathology^1^ to identification of suicidality in electronic health records (EHR) in psychiatry.^2^ While they are often highly accurate and reliable, it is rarely reported in detail how the design choices that led to the presented results were made - be it by training a foundation model from scratch, fine-tuning an existing model on relevant data, or by engineering prompts for in-context learning. Briefly, prompt engineering (PE) involves describing a task in natural language to guide an LLM in completing it.^3^ In principle, it is the most efficient solution^4^ and should be thoroughly evaluated before opting for another strategy. The relevance of structured PE is further underscored by the rapid depreciation of static rules on prompt engineering with new model releases.^5^ Recently updated guidance on prompting by Google and Anthropic,^6,7^ two makers of such models, underscore this dynamic development. Despite growing LLM use, there is no consensus on how to evaluate or structure prompts in medical applications.^2–4,8,9^ Given these challenges, a modular and testable approach is needed. To address this dilemma, we introduced clickBrick, a systematic, reproducible framework for PE in medicine, which enables direct comparison of prompt formats, including few-shot and chain-of-thought prompts,^3^ illustrated across 12 clinical information extraction tasks from unstructured medical free text. Those tasks cover the whole of psychopathology, ranging from substance use to suicidality in a transdiagnostic, holistic representation of patient behavior, cognition and emotion.^10^ clickBrick proposes a Lego-style, step-wise protocol for designing prompts that guide large language models (LLMs) in clinical work (figure 1A). The researcher begins with a naïve “zero-shot” *baseline task query* (“Does the patient show symptoms of depression?”) and then *clicks on successive prompt-bricks* that add (a) a *professional role* (“You are an experienced psychiatrist.”), (b) “one-shot” concise *operational definitions* of the diagnostic target (“Depression is characterized by sad mood, reduced energy or drive, and a loss of interest.”), (c) “few-shot” *labelled examples* of e.g. positive and negative cases (“‘The patient displays elevated mood.’ → no depressive symptom, ‘The patient felt increasingly sad and hopeless.’ → depressive symptoms present”), (d) and an explicit request for *Chain-of-Thought (CoT) reasoning* (“Think step by step!”). For a description of all relevant prompts across all 12 tasks see supplemental table S1. Finally, the designer benchmarks every prompt configuration against expert-curated ground truth, ideally verified by at least 3 blinded subject matter experts with consensus, majority, or combined method resolution^11^ - to discover the most reliable prompt stack. To scale analysis of unstructured medical text without the need for complete subject-matter expert annotation, we suggest a prediction of structurally available information, like ICD-10 diagnosis to compare most vs. least accurate prompting strategies as a demonstration of the usefulness of such systematic prompt engineering.

**Figure 1:**
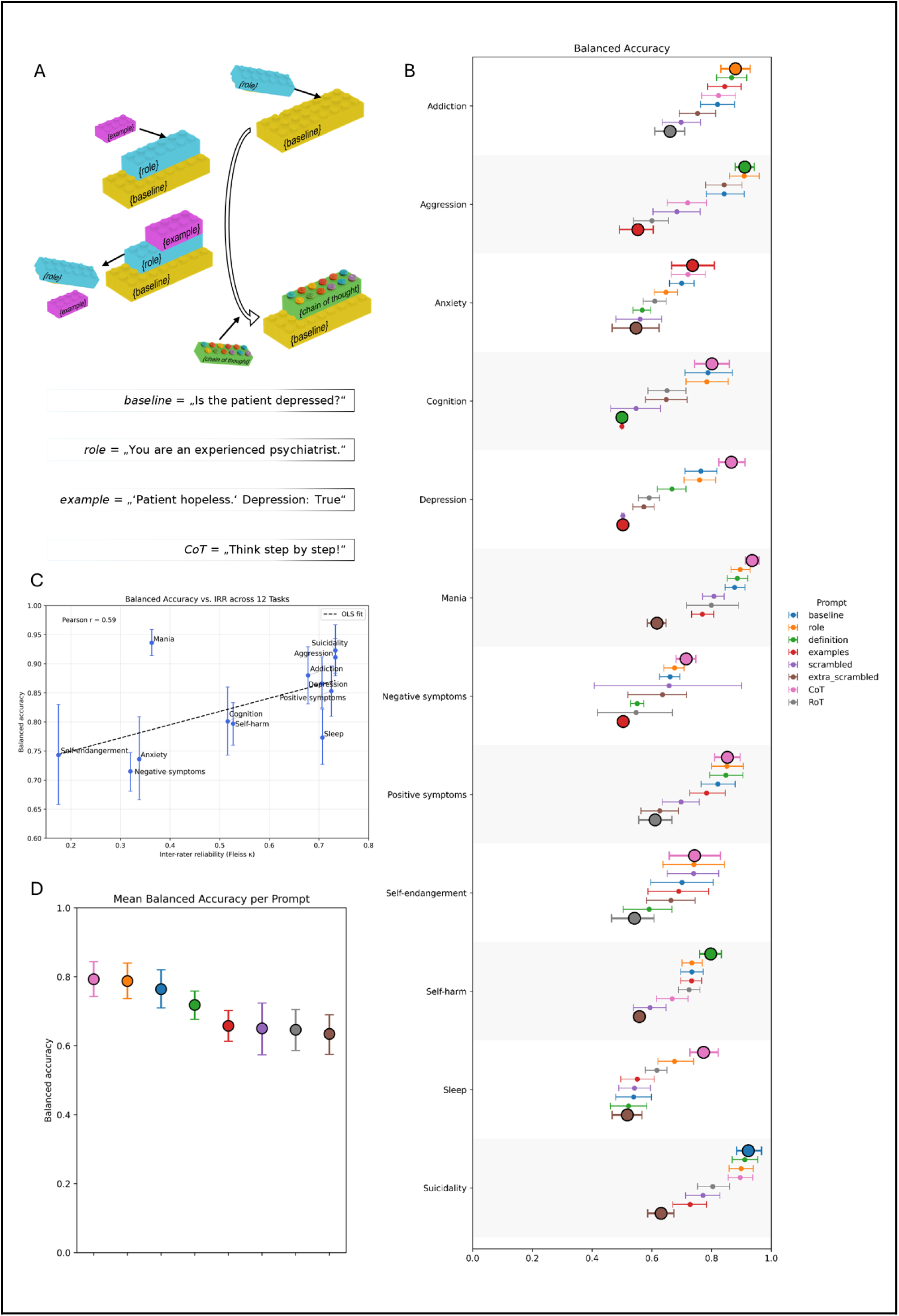
(A) ClickBrick approach: successively more structured prompts are designed. A task-defining baseline is complemented by role, example and CoT-prompts, as indicated by the colored bricks. (B) Balanced Accuracy (BAcc) for all domains showing results for all prompts, highlighting best and worst prompting strategies (bold), error bars marking 95%-CIs from 2,000-fold bootstrapping across three independent experimental runs. BAcc_Best_ is significantly higher than BAcc_worst_ with p < 0·001 for all domains. P-values were Benjamini-Hochberg-adjusted across all domains and metrics (false-discovery-rate ≤ 0·05). (C) IRR and BAccs per task are correlated with medium strength (Pearson’s r = 0·589, p = 0·044). (D) Ranking of prompt BAccs. CoT: Chain-of-Thought. RoT: Reflection-of-Thought.^3^ OLS: ordinary least squares. IRR: Inter-rater-reliability. See supplemental table S2 and supplemental figure S5 for other performance metrics.

**Figure 2:**
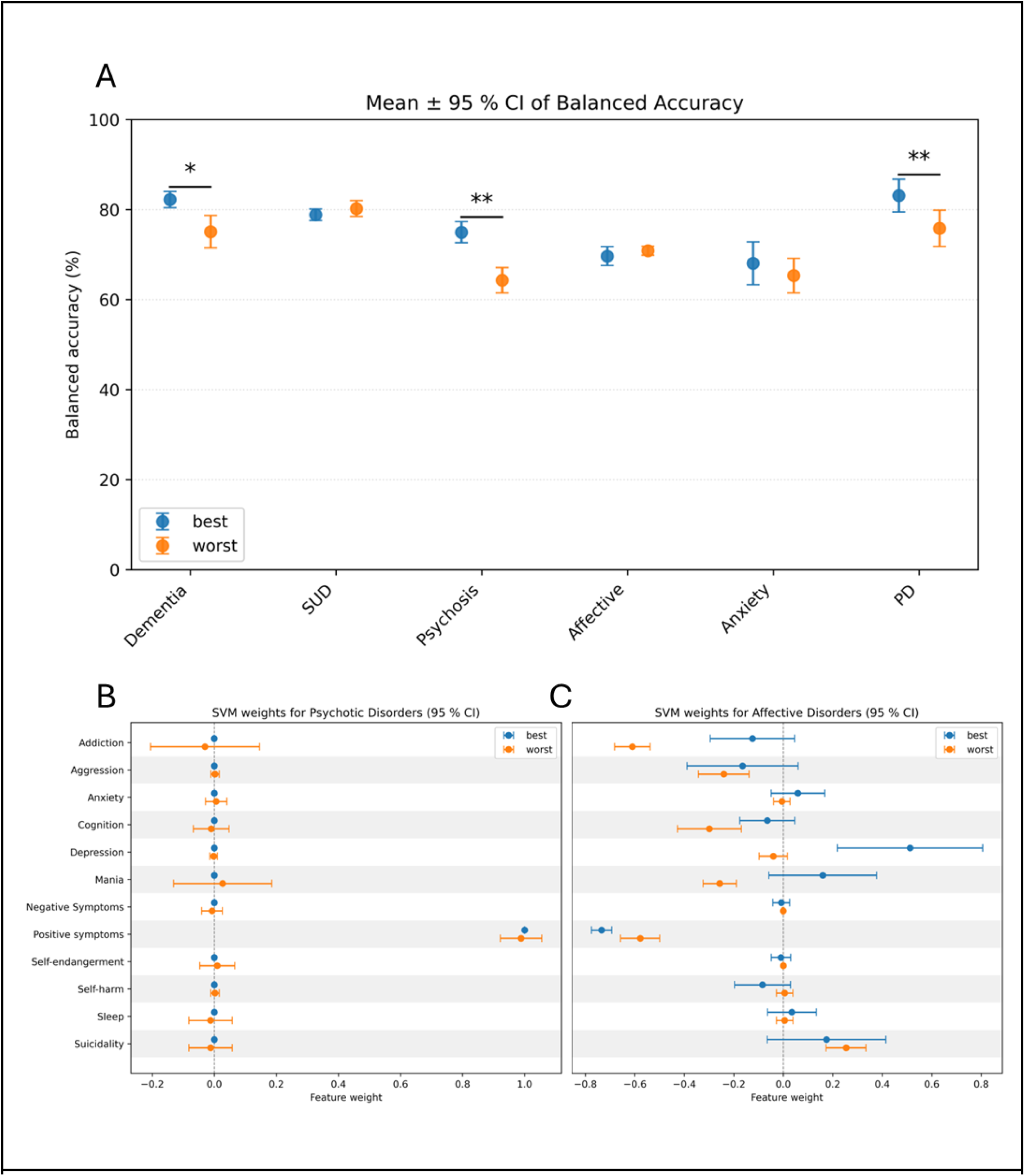
(A) BAcc achieved by SVM when trained on the least-noisy (“best”) versus the most-noisy (“worst”) version of the LLM-extracted data for each diagnosis group. “Best” (blue) and “worst” (orange) mark the mean BAcc across the 10 cross-validation folds with 95%-CI. Black horizontal bars connect paired conditions that differ significantly (two-sided Wilcoxon signed-rank test, n = 10, Benjamini-Hochberg adjusted for multiple comparisons). Significance levels indicated by asterisks above the bar (* p < 0·05, ** p < 0·01). (B) and (C) Selected, instructive feature weights (see figure S6 for complete weights): (B) SVM feature weights for F2/Psychotic Disorders: “best” and “worst” models both rely mainly on positive symptoms (i.e. hallucinations, imperative voices, etc) for classification. (C) SVM feature weights for F3/Affective Disorders: the “best” model is most influenced by the presence of depression, mania, and suicidality and the absence of positive symptoms in its classification, while the “worst” models rely equally on the absence of positive symptoms, and the presence of suicidality and is influenced more by the absence of mania, cognitive impairment or signs of addiction. See table S3 for full information.

## Methods

### Systematic Prompt Evaluation and Optimization

Pre-trained transformers exhibit a wide range of problem solving capabilities, including psychiatric classification tasks, without being programmed to do so. This ability can be harnessed via prompt engineering and in-context learning, with great difference in quality depending on this prompt.^12^ We present here a comprehensive strategy for stepwise evaluation and optimization of prompts, used for information extraction tasks, though not limited to that natural language processing application. Expert-crafted prompts are decomposed into their semantic components and their combinations subsequently tested as prompt (figure 1A) and compared, as introduced above. The clickBrick approach was also extended to a Chain-of-Thought and a simple Reflection-of-Thought prompt,^3^ for a full list of all prompts see supplemental table S1. It allows every researcher or clinician to define for each specific use case the relevant prompts and offers a versatile framework for systematic prompt engineering. Note that it does not provide a definite method of identifying the globally best prompt for any given case, only the one in the local distribution, as is discussed below. All prompts were tested with an open-weight LLM (Llama-3.1-70B-Instruct)^13^ locally run with the llama.cpp^14^ project. All research procedures were conducted in accordance with the Declaration of Helsinki. Ethics approval was granted by the ethics committee of Technical University Dresden, reference number BO-EK-400092023. Informed consent was not necessary for this study because the research involved data from which all personal identifiers had previously been removed. The design of the study ensured that there was no interaction or intervention with participants and no potential for harm or invasion of privacy.

### Definition of Transdiagnostic Information Extraction Tasks

The classic diagnostic categories for psychiatric disorders appear to group sometimes disparate entities together. A transdiagnostic approach to psychopathology aims to describe and combine traits and states in a way that might enable better description of patient presentation with potential to identify subgroups and treat them with greater precision.^15^ Here we use an adapted paradigm^10^ evaluating 12 dimensions of psychopathology: anxiety, addiction, cognition, depression, hostility, mania, negative and positive symptoms, sleep and self harm (with the dimensions self-endangerment, non-suicidal self harm, and suicidality).

### Ground Truth Curation

For a dataset of 2,520 cases from 1,692 individual patients, admission notes from the electronic health records of a large German supra-maximum psychiatric acute care ward were analyzed with a locally run and privacy preserving pipeline^16^ to automatically extract a binary evaluation (present/not present) on each of the transdiagnostic parameter from the initial medical evaluation (with the absence of reporting on the parameter being interpreted as not present). Prompts were optimized for the highest identification balanced accuracy compared against a thrice independently rated ground truth for a sub-sample of 100 cases by 3 subject-matter experts (FGV: resident psychiatrist, VM: trained research assistant, PM: attending psychiatrist), for detailed sample characteristics see Wiest and colleagues.^2^ For each of the 12 symptom domains we assessed inter-rater reliability across the three independent clinicians in two complementary ways. First, we quantified absolute (raw) agreement as the proportion of subjects whose three ratings were identical (0, 1, “missing”, or not rated). Second, chance-corrected reliability was estimated with Fleiss’ κ, appropriate for 3 raters and nominal data.

### Statistical Analysis

If not indicated otherwise, uncertainty was quantified via non-parametric bootstrapping test: 2,000 subject-level resamples with replacements were drawn, κ and BAcc respectively recomputed for each, and the 2.5^th^ and 97.5^th^ percentiles of the bootstrap distribution reported as the 95% confidence interval. We assessed the association between rater agreement and task accuracy by computing Pearson product–moment correlation coefficients for Fleiss’ κ with BAcc for all 12 domains. Additionally, the most accurate prompting strategy is compared to the least accurate one for the ensuing classification task (see below) to gauge the relevance of information extraction accuracy, with a two-sided Wilcoxon signed-rank to determine significant differences across folds. Throughout, multiple testing was addressed with Benjamini-Hochberg adjustment for a false-discovery-rate ≤ 0·05. All calculations and performance evaluations were done using pandas 2.2.3^17^, numpy 2.2.0^18^, statsmodels 0.14.4^19^, and Scikit-learn 1.6.0^20^ packages in python 3.12.3^21^. The analytic code is available at: https://github.com/verrik/clickBrick_PromptEngineering. The pipeline for local deployment of LLMs in a privacy-preserving clinical setting was described elsewhere^22,23^ and is available here: https://github.com/KatherLab/LLMAIx. Inferences with the local Llama-3.1-70B-Instruct model were run on one Nvidia A6000 GPU (48BG VRAM) on hospital premises. For an exemplary, fictional case vignette see supplemental table S1.

### Prediction of Diagnosis at Discharge Using Machine Learning

We employed a linear support vector machine to predict eventual ICD-10 psychiatric primary diagnoses (implemented in NeuroMiner 1.2^24^, MatLab 9.14.0.2206163 (R2023a)^25^, grouped by the 6 main diagnostic groups represented in our sample F0-F4 (F0 - dementias, F1 - substance use disorders, F2 - psychotic disorders, F3 - affective disorders, F4 - anxiety disorders, and F6 (personality disorders, here exclusively borderline PD), excluding the groups not represented in our study sample below a threshold of 50 cases (detailed description see supplemental figure S4). The model was trained on 2,520 cases from 1,692 individual cases using 12 binary features (0/1) reflecting key psychopathological domains (e.g., suicidality, addiction, depression), derived from clinician letters via LLM-based annotation. Classification followed a one-vs-rest strategy for six diagnostic categories (F0, F1, F2, F3, F4, F6), and for each, two models were run: one using features from the best-performing LLM prompt (based on balanced accuracy), and one using the worst. To mitigate class imbalance, ranging from 5.60% (F6), 5.79% (F4), 8.37% (F0), 22.7% (F2), 25.6% (F3), to 30.44% (F1), we applied instance-weighted hyperplane adjustment via the LIBSVM 3.1.2 implementation (C-SVC with L1-loss), ensuring minority classes received proportionally higher weighting during optimization. We did not apply oversampling or undersampling for class balancing, as such techniques have been questioned for potentially reducing model generalizability and performance^26^. We used nested 10-fold cross-validation to prevent information leakage, enable hyperparameter tuning, and ensure reliable generalization. No imputation was necessary due to complete data. The SVM regularization parameter (C) was tuned across 11 logarithmically spaced values ranging from 0.015625 to 16. No filter or wrapper-based feature selection was performed given the low-dimensional feature space relative to the sample size. The case-to-feature ratio was 210:1 (2520 cases and 12 features), supporting stable model estimation without risk of overfitting.

## Results

Overall, clickBrick increased the mean BAcc by 26·5% from 56·2% (95%-CI 53·3-59·1%) with the worst prompts to 82·8% (95%-CI 81·2-84·4%) with best. The largest increases were found for depressive symptoms (36%), mania (32%) and cognitive impairment (30%). Increase above the naïve baseline prompt was on average a more modest, but still meaningful 6·3% from 76·4% (95%-CI 75·9-77·0%) to 82·8% (95%-CI 81·2-84·4%), with largest increases for impaired sleep (23·4%), depressive symptoms (10·2%) and aggression (6·9%) (figure 1B). In accordance with the more recent advent of reasoning models, a simple CoT and a more complex RoT prompt were included in the clickBrick hierarchy. As hypothesized, CoT prompting yielded the highest accuracy in 7 out of 12 domains, and was statistically indistinguishable from the top-performing prompt in 3 additional domains after adjustment (p ≥ 0·05, table S2). However, its effectiveness varied by task: CoT was significantly outperformed by a 3-shot prompt for both aggression and self-harm (p < 0·05, table S2), underscoring the importance of domain-specific evaluation. What is more, the second reasoning prompt (RoT) that did display superior performance elsewhere^3^ showed poor accuracy in our sample, performing worst for 3 out of 12 domains. Semantic noise in the form of deliberately introduced, script-generated typing errors making the expert-designed prompts harder to interpret by the LLMs^27^ was injected in an attempt to force underperformance for the sake of this argument. Noisy prompts did perform worst in 5 out of 12 domains (1D). Inter-rater reliabilities (IRR) ranged from Fleiss’ κ of 0·18 for self-endangerment to 0·73 for aggression and suicidality, designating slight to substantial agreement^28^ (table S2). The best results for BAccs after clickBrick PE were correlated with medium strength to IRR (Pearson’s r = 0·589, p = 0·044, 1C).

To evaluate potential downstream utility and provide internal validation, we extended our clickBrick-optimized extraction to a dataset of 2,520 individual cases from 1,692 patients with a mean age of 45·2 ± 0·76 years and a balanced gender distribution (female: 1,303 cases, male: 1,215 cases, non-binary: 2 cases). Here, we used linear support vector machines (SVM) to predict final diagnosis at discharge from the clinical notes taken at admission to evaluate the optimization’s impact on potential clinical utility. We predicted class-vs-other for all relevant ICD-10 diagnostic groups that were represented in our sample (figure S4: F1 - 767 cases with substance use disorders, F3 - 645 cases with affective disorders, F2 - 572 cases with psychotic disorders, F0 - 211 cases with dementia, F4 - 146 cases with anxiety disorders, F6 - 141 cases with personality disorders - here exclusively borderline, and 38 other cases). Overall classification performance (in BAcc) increased moderately with the “best” strategy, rising from 71·9% to 76·2%. Significant improvements in classification were observed for substance use, psychotic and personality disorders, but not for the rest (p < 0·05, Benjamini-Hochberg adjusted, 2A). We subsequently analyzed feature weights to identify the clinical domains the SVM relied on most: the larger (in absolute value) the weight, the more that domain drives the decision. For psychotic disorders (2B), positive symptoms carry the top weight in both the “best prompt” and “worst prompt” models, confirming their central diagnostic importance. Yet model quality is greatly improved from a BAcc of 64% for “worst” to 75% for “best” prompting strategy, showing that upstream clickBrick, not a different leading feature, explained the performance gain. Conversely, for affective disorders the distribution of feature weights shifts substantially between models (2C). Despite this shift, classification performance remained statistically indistinguishable between the “worst” (70·8% BAcc) and “best” (69·7% BAcc) prompting strategies.

## Discussion

We show here that LLMs in clinical psychiatry offer a way to rapidly and safely aggregate and evaluate available information, e.g. from EHRs. Conversely, failure to synthesise relevant information delays diagnosis and thus treatment and often leads to considerable suffering, especially for patients with mental disorders like bipolar disorder or schizophrenia, which take between approximately 1 and 6 years to correct diagnosis.^29,30^ With clickBrick, we provide a way to optimize LLMs for the challenge of predictive modelling based on EHR to better characterize the “deep patient”.^31^ The unstructured free text thus made available encapsulates highly ecologically valid clinical data.^2,22^ We offer a starting point for future systematic evaluation of LLMs for information extraction, and in principle for any task that is defined by natural language or in fact multi-modal prompts, e.g. for pathological classification.^1^ ClickBrick addresses the currently highly heterogeneous reporting quality of prompting strategies.^2,3^ It encourages systematic and transparent iterative prompt engineering to enable better model performances. Recent evidence of sensitivity to prompt engineering even in new and large models^32^ underlines the relevance of a framework like clickBrick. Our proposed approach to PE is proven here to be a relevant means of improving the systematic evaluation of in-context learning: while we were consistently able to identify prompts for improved information extraction accuracy across all domains of interest, we could not deduct simple heuristics for the optimal prompt across all tasks. A large benefit derived indeed from identifying *under-performing* prompts that were prima-facie expected to work better, like few-shot prompting with added examples or the RoT reasoning prompt. And although the CoT reasoning prompt performed mostly superior (e.g. for anxiety), simpler prompts did better for several other psychopathological constructs, including the naïve baseline prompt for suicidality. At the same time, we could not reach a consistently high accuracy across all 12 domains, ranging from 71% (negative symptoms) to 94% (mania). The medium-strength positive correlation between inter-rater reliability and extraction accuracy offers a potential explanation for both observations: when high rater agreement denotes low classification difficulty for subject-matter experts, a higher LLM extraction accuracy can be expected, and vice versa. This assumption might be tested in future research.

Downstream classification of diagnostic groups based on “best” and “worst” extraction strategies was improved by clickBrick optimization, though not for all psychopathological domains, providing some internal validation on a larger dataset of patient cases. The observation of shifting feature weights suggests that the classifier may have relied on different information in response to reduced extraction fidelity.

In summary, we do show that comprehensive trans-diagnostic patient psychopathologies can be extracted from EHRs with good to excellent performance, which extends the existing knowledge on the capabilities of LLMs in psychiatry beyond single symptoms or social determinants of health.^2,33^ Compared to traditional natural language processing methods,^34^ there appear to be no principal limits to LLM applicability while their inclusivity using open-source resources and limited need for computer sciences experience makes them more applicable throughout different health care systems, languages and regions, including many low and middle income countries and other underserved communities.

Note that clickBrick does not provide a definitive method of identifying the globally best prompt for any given task, only the best one in the local distribution of expert-designed prompts. The merit of this approach appears to be a high consistency across tasks^35^, which is also evident in this work with high accuracies larger than 71%. Still, an alternative approach would be the automated generation and subsequent testing of prompt variations by other LLMs, e.g. based on only the initial baseline prompt, as has been described for studies with no medical background,^36^ on partly synthetic clinical data,^37^ or static medical benchmarks.^38,39^ While this will introduce other limitations (like overfitting to the ground truth sub-sample), we encourage further research into such an automated approach to clickBrick prompting, when coupled with rigorous external validation. Finally, while we did address a core ethical consideration by safeguarding data privacy with locally hosted models on hospital premises, we could not systematically assess model biases. And while we identified patient preferences for academic-led exploration of AI driven data analysis over industry research in mental health (Fesl et al, unpublished), we did not involve experts with lived experience in the development of clickBrick. We thus recommend the analysis of differential performance for subgroups^40^ based for example on gender or ethnicity for any future study that is primarily concerned with real-world applications of LLMs. We further recommend meaningful patient involvement to identify actual unmet needs that AI information extraction from EHRs might help solve, like risk stratification for the prevention of coercive measures.

## Conclusion

This study demonstrates that a systematic prompt engineering framework like clickBrick significantly improves information extraction from clinical free text with LLMs. Thus, for future research incorporating foundational generative AI models, we encourage the inclusion of a transparent reporting strategy on prompt composition, as clickBrick aims to be. Sticking to the same prompt for all domains would have led to performance reductions in information extraction in this study. Consequently, the clickBrick approach to prompting and subsequent reporting taken here might serve as a starting point for future use of natural-language-guided AI in medicine, for the sake of both the uncovering of the most efficient uses of genAI in medicine and of scientific reproducibility in this context.

## Author Contributions

FGV and PM conceptualized the study and developed the methodology in close coordination with ICW and JNK. FGV, VM and PM systematically extracted information from medical free text and determined the ground truth by consensus procedure. FGV and FH developed the scripts and ran the experiments. FGV, FH and PM accessed and verified data. FGV, FH, VM, FW, ICW and PM were writing and reviewing the initial manuscript. All authors were refining the draft. PM, ICW, AP, MB, and JNK provided supervision and resources for the project.

## Data availability statement

The source data are not publicly available because it contains real-world clinical information that could compromise the privacy of research participants.

## Analytic Code Availability

The information extraction pipeline is publicly available at https://github.com/KatherLab/LLMAIx. The analytic code is available at https://github.com/verrik/clickBrick_PromptEngineering.

## Research Material Availability

In compliance with our commitment to research transparency, we maintain accurate records of the source code associated with our manuscript, available at https://github.com/KatherLab/LLMAIx & https://github.com/verrik/clickBrick_PromptEngineering. However, the data itself is proprietary and not available for sharing in order to maintain privacy of patients. The source code necessary for replicating our procedures is openly available to other researchers.

## AI Use Statement

Artificial intelligence in the form of large language models (ChatGPT plus, o3 and others; local instances of Llama, 3.1-70B-Instruct and others) have been used to supplement the systematic literature research that this work is based on. Analytic code and feedback on the analysis plan was supported by AI, as was proof-reading for more clarity and brevity. All output was scrutinized by the authors and used only if deemed accurate.

## References

1 Ferber D, Wölflein G, Wiest IC, et al. In-context learning enables multimodal large language models to classify cancer pathology images. Nat Commun 2024; 15: 10104.

2 Wiest IC, Verhees FG, Ferber D, et al. Detection of suicidality from medical text using privacy-preserving large language models. Br J Psychiatry 2024; : 1–6.

3 Wang L, Chen X, Deng X, et al. Prompt engineering in consistency and reliability with the evidence-based guideline for LLMs. Npj Digit Med 2024; 7: 1–9.

4 Zhang X, Talukdar N, Vemulapalli S, et al. Comparison of Prompt Engineering and Fine-Tuning Strategies in Large Language Models in the Classification of Clinical Notes. AMIA Summits Transl Sci Proc 2024; 2024: 478–87.

5 Meincke L, Mollick ER, Mollick L, Shapiro D. Prompting Science Report 2: The Decreasing Value of Chain of Thought in Prompting. 2025; published online June 8. https://papers.ssrn.com/abstract=5285532 (accessed June 8, 2025).

6 Prompt engineering overview. Anthropic. https://docs.anthropic.com/en/docs/build-with-claude/prompt-engineering/overview (accessed June 2, 2025).

7 Boonstra L. Prompt Engineering. 2025; published online Feb. https://www.kaggle.com/whitepaper-prompt-engineering.

8 Hein D, Christie A, Holcomb M, et al. Prompts to Table: Specification and Iterative Refinement for Clinical Information Extraction with Large Language Models. 2025; : 2025.02.11.25322107.

9 Ganzinger M, Kunz N, Fuchs P, et al. Automated generation of discharge summaries: leveraging large language models with clinical data. Sci Rep 2025; 15: 16466.

10 Correll CU, Dombi ZB, Barabássy Á, Németh G, Brevig T, McIntyre RS. The Transdiagnostic Global Impression - Psychopathology scale (TGI-P): Initial development of a novel transdiagnostic tool for assessing, tracking, and visualising psychiatric symptom severity in everyday practice. Eur Neuropsychopharmacol J Eur Coll Neuropsychopharmacol 2024; 88: 31–9.

11 McCluskey R, Enshaei A, Hasan BAS. Finding the Ground-Truth from Multiple Labellers: Why Parameters of the Task Matter. 2021; published online Feb 16. DOI:10.48550/arXiv.2102.08482.

12 Chen B, Zhang Z, Langrené N, Zhu S. Unleashing the potential of prompt engineering in Large Language Models: a comprehensive review. 2023; published online Oct 27. DOI:10.48550/arXiv.2310.14735.

13. meta-llama/Llama-3.1-70B-Instruct · Hugging Face. 2024; published online Dec 6. https://huggingface.co/meta-llama/Llama-3.1-70B-Instruct (accessed June 26, 2025).

14 ggml-org/llama.cpp. 2025; published online June 26. https://github.com/ggml-org/llama.cpp (accessed June 26, 2025).

15 Oliver D, Chesney E, Cullen AE, et al. Exploring causal mechanisms of psychosis risk. Neurosci Biobehav Rev 2024; 162: 105699.

16 KatherLab/LLMAIx. 2025; published online April 10. https://github.com/KatherLab/LLMAIx (accessed April 12, 2025).

17 The pandas development. pandas-dev/pandas: Pandas team. 2020; published online Feb. DOI:10.5281/zenodo.3509134.

18 Harris CR, Millman KJ, Walt SJ van der, et al. Array programming with NumPy. Nature 2020; 585: 357–62.

19 Seabold S, Perktold J. statsmodels: Econometric and statistical modeling with python. In: 9th Python in Science Conference. 2010.

20 Pedregosa F, Varoquaux G, Gramfort A, et al. Scikit-learn: Machine Learning in Python. J Mach Learn Res 2011; 12: 2825–30.

21 Python 3.12 documentation. Python Doc. https://docs.python.org/3/ (accessed June 20, 2025).

22 Wiest IC, Leßmann M-E, Wolf F, et al. Deidentifying Medical Documents with Local, Privacy-Preserving Large Language Models: The LLM-Anonymizer. NEJM AI 2025; 2: AIdbp2400537.

23 Wiest IC, Wolf F, Leßmann M-E, et al. LLM-AIx: An open source pipeline for Information Extraction from unstructured medical text based on privacy preserving Large Language Models. 2024; : 2024.09.02.24312917.

24 Koutsouleris N, Kambeitz-Ilankovic L, Ruhrmann S, et al. Prediction Models of Functional Outcomes for Individuals in the Clinical High-Risk State for Psychosis or With Recent-Onset Depression: A Multimodal, Multisite Machine Learning Analysis. JAMA Psychiatry 2018; 75: 1156–72.

25 The MathWorks Inc. MATLAB version: 9.14.0.2206163 (R2023a). 2023. https://www.mathworks.com.

26 Carriero A, Luijken K, de Hond A, Moons KGM, van Calster B, van Smeden M. The Harms of Class Imbalance Corrections for Machine Learning Based Prediction Models: A Simulation Study. Stat Med 2025; 44: e10320.

27 Hughes J, Price S, Lynch A, et al. Best-of-N Jailbreaking. 2024; published online Dec 19. DOI:10.48550/arXiv.2412.03556.

28 Landis JR, Koch GG. The Measurement of Observer Agreement for Categorical Data. Biometrics 1977; 33: 159–74.

29 Drake RJ, Husain N, Marshall M, et al. Effect of delaying treatment of first-episode psychosis on symptoms and social outcomes: a longitudinal analysis and modelling study. Lancet Psychiatry 2020; 7: 602–10.

30 Scott J, Graham A, Yung A, Morgan C, Bellivier F, Etain B. A systematic review and meta-analysis of delayed help-seeking, delayed diagnosis and duration of untreated illness in bipolar disorders. Acta Psychiatr Scand 2022; 146: 389–405.

31 Miotto R, Li L, Kidd BA, Dudley JT. Deep Patient: An Unsupervised Representation to Predict the Future of Patients from the Electronic Health Records. Sci Rep 2016; 6: 26094.

32 Zhuo J, Zhang S, Fang X, Duan H, Lin D, Chen K. ProSA: Assessing and Understanding the Prompt Sensitivity of LLMs. In: Al-Onaizan Y, Bansal M, Chen Y-N, eds. Findings of the Association for Computational Linguistics: EMNLP 2024. Miami, Florida, USA: Association for Computational Linguistics, 2024: 1950–76.

33 Guevara M, Chen S, Thomas S, et al. Large language models to identify social determinants of health in electronic health records. Npj Digit Med 2024; 7: 1–14.

34 Jackson RG, Patel R, Jayatilleke N, et al. Natural language processing to extract symptoms of severe mental illness from clinical text: the Clinical Record Interactive Search Comprehensive Data Extraction (CRIS-CODE) project. BMJ Open 2017; 7: e012012.

35 Zhou Y, Zhao Y, Shumailov I, Mullins R, Gal Y. Revisiting Automated Prompting: Are We Actually Doing Better? 2023; published online June 22. DOI:10.48550/arXiv.2304.03609.

36 Yang C, Wang X, Lu Y, et al. Large Language Models as Optimizers. 2024; published online April 15. DOI:10.48550/arXiv.2309.03409.

37 Yao Z, Jaafar A, Wang B, Yang Z, Yu H. Do Physicians Know How to Prompt? The Need for Automatic Prompt Optimization Help in Clinical Note Generation. 2024; published online July 5. DOI:10.48550/arXiv.2311.09684.

38 Liu X, Ji K, Fu Y, et al. P-Tuning: Prompt Tuning Can Be Comparable to Fine-tuning Across Scales and Tasks. In: Muresan S, Nakov P, Villavicencio A, eds. Proceedings of the 60th Annual Meeting of the Association for Computational Linguistics (Volume 2: Short Papers). Dublin, Ireland: Association for Computational Linguistics, 2022: 61–8.

39 Wu S, Koo M, Scalzo F, Kurtz I. AutoMedPrompt: A New Framework for Optimizing LLM Medical Prompts Using Textual Gradients. 2025; published online Feb 21. DOI:10.48550/arXiv.2502.15944.

40 Zack T, Lehman E, Suzgun M, et al. Assessing the potential of GPT-4 to perpetuate racial and gender biases in health care: a model evaluation study. Lancet Digit Health 2024; 6: e12–22.

