## Supplemental tables S1-S3 for "clickBrick Prompt Engineering: Optimizing Large Language Model Performance in Clinical Psychiatry"

**Supplemental Table S1**: Prompts for all domains

| Prompt | Addiction | Aggression | Anxiety | Cognition | Depression | Mania | Negative symptoms | Positive symptoms | Self-endangerment | Self-harm | Sleep | Suicidality |
|---|---|---|---|---|---|---|---|---|---|---|---|---|
| **baseline (Does the patient …)** | use drugs or alcohol? | show aggressive behaviour towards others? | show symptoms of anxiety? | show cognitive impairment? | show depressive symptoms? | show manic symptoms? | show negative symptoms? | show positive symptoms? | engage in behaviour that endangers themselves? | exhibit self-harming behaviour? | have sleep problems? | exhibit suicidal behaviour? |
| **role** | You are an experienced, attentive physician with specialized knowledge in psychiatry. | | | | | | | | | | | |
| **definition** | Any recent substance use (alcohol, cannabis, meth, cocaine, heroin, ketamine, etc.). Nicotine and caffeine not counted. | Hostile, irritable, explosive, insulting, or deliberate harm towards others. | Symptoms include somatic symptoms (palpitations, chest pain, shortness of breath, dizziness, muscle tension), nervousness, free-floating anxiety, depersonalization, derealization, fear of dying, fear of losing control, phobias. | Reduced comprehension, concentration, memory deficits. | Defined by ICD-10: low mood, lack of affect, reduced emotional responsiveness, reduced motivation/activity, inability to feel pleasure, loss of interest, fatigue, poor concentration, low self-esteem, guilt, feelings of worthlessness, agitation, loss of libido. | Defined by ICD-10: disproportionate elevated mood, increased drive, heightened sense of vitality, sociability, libido, overconfidence, reduced need for sleep, recklessness, irritability, reduced concentration, logorrhea, delusions of grandeur, hallucinations. | Loss of cognitive/psychomotor abilities: anhedonia, apathy, blunted affect, attention problems, social withdrawal, alogia, psychomotor retardation. | Defined as: ego disturbances (thought insertion, withdrawal, broadcasting), perceptual disturbances/ hallucinations (auditory, visual, olfactory, tactile), delusions, formal thought disorder (loosening of associations, blocking, neologisms), bizarre behavior. | Behavior (conscious or unconscious) placing oneself at risk of harm. | Intentional, often repetitive damage to one's own body without suicidal intent. Superficial cuts/scars are typical. | Sleep onset problems, maintenance problems, early waking, poor sleep quality, daytime fatigue, excessive sleepiness. If well-controlled on medication, considered not present. | Thoughts of suicide, planning suicide, recent suicide attempts, or omission of life-saving actions with intent to die. |
| **examples** | Example 1: Occasional cannabis and meth use → Yes. Example 2: Denies alcohol and drug use → No. Example 3: Admitted intoxicated → Yes. | Example 1: Aggressive and hard to limit → Yes. Example 2: Cannot rule out aggression → Yes. Example 3: No signs of aggression → No. | Example 1: No anxiety or compulsions → No. Example 2: Fear of failure and disappointing family → Yes. Example 3: Suspicious, anxious, nervous → Yes. | Example 1: Slightly reduced concentration → Yes. Example 2: Fully oriented, no deficits → No. Example 3: Slightly impaired after alcohol intoxication → Yes. | Example 1: Patient is withdrawn, unresponsive → Yes. Example 2: Friendly and outgoing → No. Example 3: Mood appropriate → No. | Example 1: Patient is irritable, elevated mood → Yes. Example 2: Increased drive, confrontational → Yes. Example 3: Patient friendly and calm → No. | Example 1: Hardly leaves home, reduced family contact → Yes. Example 2: Psychomotor retardation, apathy → Yes. Example 3: Open and responsive → No. | Example 1: Patient reports hearing voices and delusions → Yes. Example 2: Acute intoxication with psychotic experience → Yes. Example 3: No hallucinations or delusions → No. | Example 1: Acutely endangered due to misperception → Yes. Example 2: No signs of self-endangerment → No. Example 3: Found disoriented in traffic → Yes. | Example 1: Self-harm scars present → Yes. Example 2: Skin intact → No. Example 3: Recently self-harmed due to stress → Yes. | Example 1: Poor sleep, frequent waking → Yes. Example 2: No sleep problems → No. Example 3: Controlled with medication → No. | Example 1: Tried to jump from bridge → Yes. Example 2: Expresses wish to die → Yes. Example 3: No suicidal thoughts → No. |
| **scrambled** | DEOS ThE PATIEnt uSE drgUs OR | dOEs ThE pIATENT SOhW | does THE PaTIENt SHOW | dEos tHE PIETNaT shOW | dOES The PnIAett sHOw DISPrvESEE | deOS THE PaTIenT sHOW | dOes THe PATIENt SoHw | DOES ThE PNiATEt ShoW | dOEs The PaTEINT EnGAGE iN | dOEs tHE pENAiTt EbXHIIT | does THE PnEitAT HAvE SLEEP | dEos THe paTnEIT eBiIxHT |

| | AcOlhLO? | aGgRESSiVe bAEoVHIR TaWDRos otHERs? | SyMpTOMs oF ANXieTY? | CIGtOvNIE InmTEIRaPM ? | sYmptOMS? | MANIC SyMPTOMS? | neGaTiVE SYMPToMs? | PtIvSOiE sMMtYPOS? | BAiOhvEr THaT endANgERS TelmESSEVH ? | SHNAErI-Fm LG bveaRoih | PRoBlEmS? | siulaCdL bERVHaIO? |
|---|---|---|---|---|---|---|---|---|---|---|---|---|
| **extra scrambled (i.e., +role)** | yoU ARE AN eDEIEXNEPCR, ATTeNTiVE PyhsSCCin WiTh SALEiEZCiPD kLeGWnDOE in PSyChIATry. | | | | | | | | | | | |
| **CoT** | Think step by step! | | | | | | | | | | | |
| **RoT** | Simulate the thought process of three independent psychiatric experts analyzing the given report. Each expert should follow a step-by-step reasoning process from Step 1 to Step X and answer the question independently. After completing their individual analyses, the experts should engage in a discussion, review each other's thought processes, and collaboratively reach a final consensus. Clearly present the individual thought process of each expert, followed by the group discussion and the final conclusion. | | | | | | | | | | | |
| **case vignette** | Patient phoned the regional crisis line after escalating suicidal ideation with a specific plan (overdose on prescribed venlafaxine). Crisis clinician coordinated EMS transport. On scene, patient was tearful, expressed hopelessness, but denied attempts that day. Collateral from friend corroborates 2-week functional decline, social withdrawal, and missed workdays. Prior Psychiatric Treatment & Course: Out-patient: Diagnosed with Major Depressive Disorder (recurrent, moderate) at age 25; managed by GP with sertraline 50 mg daily (poor adherence in past 3 months). Psychotherapy: Six CBT sessions (2019) – self-terminated due to scheduling conflicts. Hospitalizations: None. Substance Use: Social alcohol (1–2 drinks/week); denies illicit substances or tobacco. Medical History: Hypothyroidism (on levothyroxine 75 µg daily, TSH last month 2.1 mIU/L); no surgeries. Family & Social History Family Psychiatric Hx: Mother with recurrent MDD; paternal uncle died by suicide (age 48). No bipolar or psychotic disorders reported. Developmental / Education: Unremarkable childhood, university degree in graphic design. Occupational: Full-time freelance designer; workload recently decreased. Living Situation: Rents one-bedroom apartment; single, limited social support. Protective Factors: Strong attachment to pet dog, stated desire to “not hurt my mother,” insight into illness, willingness to engage in treatment. Mental Status & Psychopathology Assessment. Appearance: Thin, casually dressed, fair hygiene, slumped posture. Behavior: Psychomotor retardation; cooperative, maintains eye contact. Speech: Soft, slow, decreased spontaneity. Mood: “Empty, exhausted”. Affect: Constricted, congruent with mood. Thought Process: Linear, goal-directed. Thought Content: Worthlessness, excessive guilt, passive SI progressing to active SI with plan; no homicidal ideation, delusions, or obsessions. Perception: No hallucinations. Cognition: Alert & oriented ×4; attention intact; immediate recall 3/3, 3/3 after 10 minutes. Insight/Judgment: Good insight into depressive symptoms; judgment impaired by hopelessness around future. Physical Examination (Admission): Vital Signs: BP 118/72 mmHg, HR 76 bpm, Temp 36.7 °C, RR 14 /min, $SpO_2$ 98 % RA. General: Appears stated age, no acute distress. Pupils equal, round, reactive; TMs intact; oropharynx moist. Cardiovascular/Respiratory: normal sounds( S1/2), no murmurs; lungs clear bilaterally. Abdomen: Soft, non-tender, normoactive bowel sounds. Neuro: Cranial nerves I–XII intact; motor 5/5; reflexes 2+ symmetric; gait normal. Skin: No rashes, track marks, or self-harm scars. | | | | | | | | | | | |

A string would be handed to the large language model, comprising the case report and the prompt (“{report} {prompt}”) as one continuous text. All prompts were expert-designed to give the model relevant knowledge that a human rater would benefit from. Definitions are based on ICD-10[1] and AMDP[2] manuals. Intentional noise was introduced for the two scrambled prompts in the expectation of reduced performance, based on a script to experimentally circumvent proprietary model guardrails[3]. Chain-of-Thought (CoT) and Reflection-of-Thought (RoT) prompt structures were adapted from Wang and colleagues[4]. A typical, though fictional case vignette is also shown here.

---

[1] World Health Organization, *ICD-10 : international statistical classification of diseases and related health problems : tenth revision* (World Health Organization, 2004), https://iris.who.int/handle/10665/42980.
[2] Arbeitsgemeinschaft für Methodik und Dokumentation in der Psychiatrie (AMDP), *Das AMDP-System: Manual Zur Dokumentation Des Psychischen Befundes in Psychiatrie, Psychotherapie Und Psychosomatik*, 11. vollständig überarbeitete Auflage (Göttingen: Hogrefe Verlag, 2023), https://www.hogrefe.com/de/shop/default-name-96607.html.
[3] John Hughes et al., “Best-of-N Jailbreaking” (arXiv, December 19, 2024), https://doi.org/10.48550/arXiv.2412.03556.
[4] Li Wang et al., “Prompt Engineering in Consistency and Reliability with the Evidence-Based Guideline for LLMs,” *Npj Digital Medicine* 7, no. 1 (February 20, 2024): 1–9, https://doi.org/10.1038/s41746-024-01029-4.

**Supplemental Table S2**: Performance metrics and significance levels for all domain extractions

| Condition | Prompt | Accuracy | 95% CI | BAcc | 95% CI | Precision | 95% CI | Recall | 95% CI | F1 | 95% CI | significant ΔBAcc vs· best prompt | p-value | IRR Fleiss' κ (95% CI) |
|---|---|---|---|---|---|---|---|---|---|---|---|---|---|---|
| Addiction | 1 | 0·82 | 0·04 | 0·82 | 0·04 | 0·73 | 0·07 | 0·83 | 0·08 | 0·78 | 0·04 | yes | 0·018 | 0·678 (0·577-0·774) |
| | 2 | 0·89 | 0·04 | 0·88 | 0·04 | 0·85 | 0·07 | 0·85 | 0·04 | 0·85 | 0·05 | | | |
| | 3 | 0·87 | 0·04 | 0·87 | 0·03 | 0·80 | 0·07 | 0·87 | 0·00 | 0·83 | 0·04 | | | |
| | 4 | 0·86 | 0·03 | 0·84 | 0·02 | 0·86 | 0·07 | 0·76 | 0·00 | 0·81 | 0·03 | | | |
| | 5 | 0·75 | 0·00 | 0·70 | 0·01 | 0·78 | 0·03 | 0·48 | 0·04 | 0·59 | 0·02 | | | |
| | 6 | 0·79 | 0·04 | 0·75 | 0·02 | 0·78 | 0·15 | 0·61 | 0·08 | 0·69 | 0·02 | | | |
| | 7 | 0·82 | 0·04 | 0·82 | 0·06 | 0·73 | 0·01 | 0·83 | 0·15 | 0·78 | 0·07 | no | 0·106 | |
| | 8 | 0·69 | 0·19 | 0·66 | 0·21 | 0·59 | 0·27 | 0·55 | 0·28 | 0·57 | 0·27 | yes | <0·001 | |
| Aggression | 1 | 0·86 | 0·01 | 0·84 | 0·01 | 0·70 | 0·04 | 0·80 | 0·00 | 0·75 | 0·02 | no | 0·073 | 0·733 (0·603-0·844) |
| | 2 | 0·93 | 0·02 | 0·91 | 0·07 | 0·86 | 0·03 | 0·87 | 0·15 | 0·86 | 0·06 | | | |
| | 3 | 0·87 | 0·01 | 0·91 | 0·01 | 0·65 | 0·02 | 1·00 | 0·00 | 0·79 | 0·02 | | | |
| | 4 | 0·77 | 0·00 | 0·55 | 0·00 | 0·75 | 0·00 | 0·12 | 0·00 | 0·21 | 0·00 | yes | <0·001 | |
| | 5 | 0·79 | 0·01 | 0·68 | 0·03 | 0·61 | 0·04 | 0·47 | 0·06 | 0·53 | 0·04 | | | |
| | 6 | 0·86 | 0·03 | 0·84 | 0·05 | 0·68 | 0·05 | 0·81 | 0·11 | 0·74 | 0·06 | | | |
| | 7 | 0·84 | 0·02 | 0·72 | 0·04 | 0·80 | 0·12 | 0·48 | 0·10 | 0·60 | 0·08 | yes | <0·001 | |
| | 8 | 0·80 | 0·02 | 0·60 | 0·05 | 1·00 | 0·00 | 0·20 | 0·10 | 0·33 | 0·14 | | | |
| Anxiety | 1 | 0·59 | 0·01 | 0·70 | 0·01 | 0·43 | 0·01 | 1·00 | 0·00 | 0·60 | 0·01 | no | 0·517 | 0·338 (0·239-0·438) |
| | 2 | 0·51 | 0·01 | 0·65 | 0·01 | 0·39 | 0·01 | 1·00 | 0·00 | 0·56 | 0·01 | | | |
| | 3 | 0·40 | 0·05 | 0·57 | 0·04 | 0·34 | 0·02 | 1·00 | 0·00 | 0·51 | 0·02 | | | |
| | 4 | 0·73 | 0·04 | 0·74 | 0·03 | 0·55 | 0·05 | 0·74 | 0·00 | 0·63 | 0·03 | | | |
| | 5 | 0·57 | 0·06 | 0·56 | 0·07 | 0·37 | 0·07 | 0·53 | 0·12 | 0·43 | 0·09 | | | |
| | 6 | 0·51 | 0·05 | 0·55 | 0·06 | 0·34 | 0·04 | 0·65 | 0·08 | 0·45 | 0·06 | yes | <0·001 | |
| | 7 | 0·66 | 0·05 | 0·72 | 0·03 | 0·48 | 0·04 | 0·87 | 0·08 | 0·62 | 0·03 | no | 0·806 | |
| | 8 | 0·49 | 0·08 | 0·61 | 0·03 | 0·37 | 0·03 | 0·94 | 0·08 | 0·53 | 0·02 | | | |

| | | | | | | | | | | | | | | |
|---|---|---|---|---|---|---|---|---|---|---|---|---|---|---|
| **Cognition** | 1 | 0·83 | 0·00 | 0·79 | 0·00 | 0·90 | 0·00 | 0·87 | 0·00 | 0·89 | 0·00 | no | 0·796 | 0·516 (0·352-0·656) |
| | 2 | 0·82 | 0·06 | 0·78 | 0·05 | 0·90 | 0·03 | 0·86 | 0·08 | 0·88 | 0·04 | | | |
| | 3 | 0·76 | 0·00 | 0·50 | 0·00 | 0·76 | 0·00 | 1·00 | 0·00 | 0·86 | 0·00 | yes | <0·001 | |
| | 4 | 0·76 | 0·00 | 0·50 | 0·00 | 0·76 | 0·00 | 1·00 | 0·00 | 0·86 | 0·00 | | | |
| | 5 | 0·64 | 0·03 | 0·55 | 0·05 | 0·78 | 0·03 | 0·72 | 0·02 | 0·75 | 0·02 | | | |
| | 6 | 0·80 | 0·01 | 0·65 | 0·05 | 0·82 | 0·03 | 0·93 | 0·06 | 0·87 | 0·01 | | | |
| | 7 | 0·82 | 0·05 | 0·80 | 0·07 | 0·92 | 0·04 | 0·84 | 0·04 | 0·88 | 0·03 | n.a. | n.a. | |
| | 8 | 0·77 | 0·06 | 0·65 | 0·10 | 0·83 | 0·05 | 0·89 | 0·05 | 0·86 | 0·04 | | | |
| **Depression** | 1 | 0·77 | 0·01 | 0·76 | 0·01 | 0·70 | 0·01 | 1·00 | 0·00 | 0·82 | 0·01 | yes | <0·001 | 0·706 (0·586-0·812) |
| | 2 | 0·77 | 0·02 | 0·76 | 0·03 | 0·69 | 0·02 | 1·00 | 0·00 | 0·82 | 0·02 | | | |
| | 3 | 0·68 | 0·02 | 0·67 | 0·03 | 0·62 | 0·02 | 1·00 | 0·00 | 0·76 | 0·01 | | | |
| | 4 | 0·52 | 0·01 | 0·50 | 0·01 | 0·52 | 0·01 | 1·00 | 0·00 | 0·69 | 0·01 | yes | <0·001 | |
| | 5 | 0·52 | 0·01 | 0·50 | 0·01 | 0·52 | 0·01 | 1·00 | 0·00 | 0·69 | 0·01 | | | |
| | 6 | 0·59 | 0·04 | 0·57 | 0·04 | 0·56 | 0·03 | 1·00 | 0·00 | 0·72 | 0·02 | | | |
| | 7 | 0·87 | 0·07 | 0·87 | 0·08 | 0·81 | 0·09 | 0·97 | 0·03 | 0·89 | 0·06 | n.a. | n.a. | |
| | 8 | 0·61 | 0·08 | 0·59 | 0·08 | 0·57 | 0·05 | 0·98 | 0·00 | 0·72 | 0·04 | | | |
| **Mania** | 1 | 0·78 | 0·01 | 0·88 | 0·01 | 0·29 | 0·01 | 1·00 | 0·00 | 0·45 | 0·02 | yes | <0·001 | 0·363 (0·122-0·556) |
| | 2 | 0·81 | 0·02 | 0·90 | 0·01 | 0·32 | 0·03 | 1·00 | 0·00 | 0·49 | 0·03 | | | |
| | 3 | 0·79 | 0·01 | 0·89 | 0·01 | 0·30 | 0·01 | 1·00 | 0·00 | 0·47 | 0·02 | | | |
| | 4 | 0·58 | 0·02 | 0·77 | 0·01 | 0·18 | 0·01 | 1·00 | 0·00 | 0·30 | 0·01 | | | |
| | 5 | 0·65 | 0·09 | 0·81 | 0·05 | 0·21 | 0·04 | 1·00 | 0·00 | 0·34 | 0·06 | | | |
| | 6 | 0·30 | 0·03 | 0·62 | 0·02 | 0·11 | 0·00 | 1·00 | 0·00 | 0·21 | 0·01 | yes | <0·001 | |
| | 7 | 0·88 | 0·03 | 0·94 | 0·02 | 0·44 | 0·06 | 1·00 | 0·00 | 0·61 | 0·06 | n.a. | n.a. | |
| | 8 | 0·79 | 0·06 | 0·80 | 0·09 | 0·27 | 0·07 | 0·81 | 0·16 | 0·41 | 0·09 | | | |
| **Negative symptoms** | 1 | 0·36 | 0·01 | 0·66 | 0·01 | 0·09 | 0·00 | 1·00 | 0·00 | 0·16 | 0·00 | yes | 0·016 | 0·32 (0·066-0·542) |
| | 2 | 0·39 | 0·02 | 0·68 | 0·01 | 0·09 | 0·00 | 1·00 | 0·00 | 0·16 | 0·01 | | | |
| | 3 | 0·16 | 0·04 | 0·55 | 0·02 | 0·07 | 0·00 | 1·00 | 0·00 | 0·12 | 0·00 | | | |

| | | | | | | | | | | | | | | |
|---|---|---|---|---|---|---|---|---|---|---|---|---|---|---|
| | 4 | 0·07 | 0·01 | 0·50 | 0·01 | 0·06 | 0·00 | 1·00 | 0·00 | 0·11 | 0·00 | yes | <0·001 | |
| | 5 | 0·80 | 0·03 | 0·66 | 0·02 | 0·15 | 0·02 | 0·50 | 0·00 | 0·23 | 0·03 | | | |
| | 6 | 0·41 | 0·04 | 0·64 | 0·11 | 0·08 | 0·02 | 0·89 | 0·24 | 0·15 | 0·03 | | | |
| | 7 | 0·46 | 0·03 | 0·71 | 0·02 | 0·10 | 0·00 | 1·00 | 0·00 | 0·18 | 0·01 | n.a. | n.a. | |
| | 8 | 0·34 | 0·03 | 0·55 | 0·21 | 0·07 | 0·03 | 0·78 | 0·48 | 0·12 | 0·06 | | | |
| **Positive symptoms** | 1 | 0·82 | 0·02 | 0·82 | 0·02 | 0·79 | 0·04 | 0·84 | 0·03 | 0·81 | 0·02 | no | 0·333 | 0·725 (0·614-0·826) |
| | 2 | 0·85 | 0·04 | 0·85 | 0·04 | 0·79 | 0·05 | 0·91 | 0·00 | 0·85 | 0·03 | | | |
| | 3 | 0·85 | 0·00 | 0·85 | 0·00 | 0·86 | 0·00 | 0·81 | 0·00 | 0·84 | 0·00 | | | |
| | 4 | 0·78 | 0·04 | 0·78 | 0·04 | 0·71 | 0·04 | 0·89 | 0·00 | 0·79 | 0·03 | | | |
| | 5 | 0·69 | 0·03 | 0·70 | 0·03 | 0·64 | 0·02 | 0·77 | 0·06 | 0·70 | 0·04 | | | |
| | 6 | 0·61 | 0·08 | 0·63 | 0·08 | 0·56 | 0·05 | 0·83 | 0·14 | 0·67 | 0·08 | | | |
| | 7 | 0·85 | 0·09 | 0·85 | 0·09 | 0·84 | 0·11 | 0·84 | 0·08 | 0·84 | 0·09 | n.a. | n.a. | |
| | 8 | 0·62 | 0·11 | 0·61 | 0·10 | 0·63 | 0·19 | 0·47 | 0·09 | 0·54 | 0·10 | yes | <0·001 | |
| **Self-endangerm ent** | 1 | 0·87 | 0·01 | 0·70 | 0·03 | 0·90 | 0·01 | 0·94 | 0·03 | 0·92 | 0·01 | no | 0·281 | 0·175 (0·015-0·329) |
| | 2 | 0·82 | 0·00 | 0·74 | 0·00 | 0·92 | 0·00 | 0·86 | 0·00 | 0·89 | 0·00 | | | |
| | 3 | 0·84 | 0·03 | 0·59 | 0·09 | 0·87 | 0·03 | 0·95 | 0·00 | 0·91 | 0·01 | | | |
| | 4 | 0·78 | 0·03 | 0·69 | 0·02 | 0·91 | 0·00 | 0·82 | 0·03 | 0·86 | 0·02 | | | |
| | 5 | 0·75 | 0·05 | 0·74 | 0·06 | 0·94 | 0·02 | 0·75 | 0·05 | 0·83 | 0·04 | | | |
| | 6 | 0·79 | 0·02 | 0·66 | 0·10 | 0·90 | 0·04 | 0·85 | 0·05 | 0·87 | 0·02 | | | |
| | 7 | 0·82 | 0·08 | 0·74 | 0·10 | 0·92 | 0·03 | 0·86 | 0·07 | 0·89 | 0·05 | n.a. | n.a. | |
| | 8 | 0·78 | 0·10 | 0·54 | 0·17 | 0·85 | 0·05 | 0·90 | 0·07 | 0·87 | 0·06 | yes | <0·001 | |
| **Self-harm** | 1 | 0·54 | 0·01 | 0·73 | 0·01 | 0·23 | 0·01 | 1·00 | 0·00 | 0·38 | 0·01 | yes | <0·001 | 0·527 (0·335-0·688) |
| | 2 | 0·54 | 0·14 | 0·73 | 0·08 | 0·24 | 0·05 | 1·00 | 0·00 | 0·38 | 0·07 | | | |
| | 3 | 0·65 | 0·05 | 0·80 | 0·03 | 0·29 | 0·03 | 1·00 | 0·00 | 0·44 | 0·04 | | | |
| | 4 | 0·54 | 0·17 | 0·73 | 0·10 | 0·24 | 0·07 | 1·00 | 0·00 | 0·38 | 0·09 | | | |
| | 5 | 0·37 | 0·11 | 0·59 | 0·07 | 0·17 | 0·02 | 0·90 | 0·20 | 0·29 | 0·04 | | | |
| | 6 | 0·24 | 0·32 | 0·56 | 0·19 | 0·16 | 0·06 | 1·00 | 0·00 | 0·27 | 0·09 | yes | <0·001 | |

| | | | | | | | | | | | | | | |
|---|---|---|---|---|---|---|---|---|---|---|---|---|---|---|
| | 7 | 0·48 | 0·02 | 0·67 | 0·07 | 0·20 | 0·02 | 0·93 | 0·18 | 0·33 | 0·04 | yes | <0·001 | |
| | 8 | 0·54 | 0·17 | 0·72 | 0·11 | 0·23 | 0·07 | 0·98 | 0·10 | 0·38 | 0·09 | | | |
| **Sleep** | 1 | 0·53 | 0·04 | 0·54 | 0·04 | 0·51 | 0·02 | 0·82 | 0·03 | 0·63 | 0·03 | yes | <0·001 | 0·707 (0·609-0·794) |
| | 2 | 0·67 | 0·09 | 0·68 | 0·09 | 0·63 | 0·07 | 0·80 | 0·11 | 0·71 | 0·09 | | | |
| | 3 | 0·52 | 0·10 | 0·52 | 0·10 | 0·50 | 0·07 | 0·79 | 0·08 | 0·62 | 0·07 | | | |
| | 4 | 0·55 | 0·41 | 0·55 | 0·40 | 0·58 | 0·38 | 0·58 | 0·08 | 0·57 | 0·18 | | | |
| | 5 | 0·54 | 0·02 | 0·54 | 0·04 | 0·53 | 0·02 | 0·62 | 0·65 | 0·55 | 0·24 | | | |
| | 6 | 0·52 | 0·01 | 0·52 | 0·02 | 0·51 | 0·01 | 0·58 | 0·47 | 0·53 | 0·19 | yes | <0·001 | |
| | 7 | 0·77 | 0·07 | 0·77 | 0·07 | 0·70 | 0·06 | 0·93 | 0·08 | 0·80 | 0·07 | n.a. | n.a. | |
| | 8 | 0·61 | 0·09 | 0·62 | 0·09 | 0·56 | 0·05 | 0·96 | 0·05 | 0·71 | 0·06 | | | |
| **Suicidality** | 1 | 0·93 | 0·00 | 0·92 | 0·00 | 0·92 | 0·00 | 0·97 | 0·00 | 0·94 | 0·00 | n.a. | n.a. | 0·733 (0·629-0·828) |
| | 2 | 0·91 | 0·04 | 0·90 | 0·05 | 0·90 | 0·08 | 0·95 | 0·04 | 0·92 | 0·03 | | | |
| | 3 | 0·92 | 0·01 | 0·91 | 0·01 | 0·89 | 0·00 | 0·99 | 0·02 | 0·94 | 0·01 | | | |
| | 4 | 0·75 | 0·11 | 0·73 | 0·12 | 0·74 | 0·09 | 0·89 | 0·07 | 0·81 | 0·08 | | | |
| | 5 | 0·78 | 0·06 | 0·77 | 0·05 | 0·79 | 0·02 | 0·85 | 0·10 | 0·82 | 0·06 | | | |
| | 6 | 0·69 | 0·01 | 0·63 | 0·03 | 0·65 | 0·02 | 0·98 | 0·07 | 0·78 | 0·01 | yes | <0·001 | |
| | 7 | 0·90 | 0·07 | 0·90 | 0·07 | 0·91 | 0·05 | 0·92 | 0·07 | 0·91 | 0·06 | no | 0·252 | |
| | 8 | 0·82 | 0·05 | 0·80 | 0·06 | 0·80 | 0·05 | 0·93 | 0·05 | 0·86 | 0·04 | | | |

Prompt 1: baseline, 2: role+baseline, 3: role+definition+baseline, 4: role+definition+examples+baseline, 5: scrambled (baseline), 6: extra scrambled (role+baseline), 7: Chain-of-Thought (CoT), 8: Reflection-of-Thought. BAcc - Balanced accuracy. IRR - Inter-rater reliability. Best (green) and worst (red) BAcc highlighted for each domain. Means, 95%-CIs and p-values derived from 2,000 bootstrapping iterations. P-values were Benjamini-Hochberg-adjusted across all domains and metrics (false-discovery-rate ≤ 0·05). Significance of BAcc-differences between the best and worst prompt, between the best and baseline prompt, and between the best and CoT prompt are given in the respective row, where applicable. Fleiss κ ranges: 0 - 0·20 → slight, 0·21 - 0·40 → fair, 0·41 - 0·60 → moderate, 0·61 - 0·80 → substantial, 0·81 - 1 → almost perfect.

**Supplemental Table S3**: Performance metrics, significance levels and feature weights for diagnostic group classifiers

| classifier | Dementia (F0) | | | | SUD (F1) | | | | Psychotic Disorders (F2) | | | | Affective Disorders (F3) | | | | Anxiety Disorders (F4) | | | | Personality Disorders (F5) | | | |
|---|---|---|---|---|---|---|---|---|---|---|---|---|---|---|---|---|---|---|---|---|---|---|---|---|
| mean BACC | 82.25 | | 75.07 | | 78.86 | | 80.23 | | 74.97 | | 64.31 | | 69.67 | | 70.85 | | 68.05 | | 65.34 | | 83.11 | | 75.84 | |
| 95% CI | 1.81 | | 3.61 | | 1.27 | | 1.76 | | 2.35 | | 2.80 | | 2.09 | | 0.98 | | 4.76 | | 3.84 | | 3.66 | | 4.02 | |
| p value | 0.004 | | | | 0.046 | | | | <0.001 | | | | 0.247 | | | | 0.340 | | | | <0.001 | | | |
| | Feature weights (mean, S.E.) | | | | | | | | | | | | | | | | | | | | | | | |
| Addiction | -0.54 | 0.01 | -0.63 | 0.06 | 1.00 | 0.00 | 1.00 | 0.00 | 0.00 | 0.00 | -0.03 | 0.09 | -0.12 | 0.09 | -0.61 | 0.04 | -0.48 | 0.05 | -0.55 | 0.06 | 0.00 | 0.00 | -0.03 | 0.09 |
| Aggression | 0.31 | 0.01 | 0.15 | 0.09 | 0.00 | 0.00 | 0.00 | 0.00 | 0.00 | 0.00 | 0.00 | 0.01 | -0.16 | 0.11 | -0.24 | 0.05 | -0.31 | 0.08 | -0.15 | 0.16 | 0.00 | 0.00 | 0.00 | 0.00 |
| Anxiety | -0.28 | 0.01 | -0.03 | 0.03 | 0.00 | 0.00 | 0.00 | 0.00 | 0.00 | 0.00 | 0.01 | 0.02 | 0.06 | 0.06 | -0.01 | 0.02 | 0.17 | 0.03 | 0.00 | 0.00 | 0.00 | 0.00 | 0.44 | 0.13 |
| Cognition | 0.56 | 0.01 | 0.62 | 0.04 | 0.00 | 0.00 | 0.00 | 0.00 | 0.00 | 0.00 | -0.01 | 0.03 | -0.06 | 0.06 | -0.30 | 0.07 | -0.19 | 0.06 | -0.06 | 0.13 | 0.00 | 0.00 | -0.49 | 0.06 |
| Depression | -0.25 | 0.01 | -0.06 | 0.03 | 0.00 | 0.00 | 0.00 | 0.00 | 0.00 | 0.00 | 0.00 | 0.01 | 0.51 | 0.15 | -0.04 | 0.03 | 0.12 | 0.05 | 0.42 | 0.12 | 0.00 | 0.00 | 0.45 | 0.13 |
| Mania | -0.01 | 0.01 | 0.06 | 0.03 | 0.00 | 0.00 | 0.00 | 0.00 | 0.00 | 0.00 | 0.03 | 0.08 | 0.16 | 0.11 | -0.26 | 0.03 | -0.02 | 0.10 | -0.06 | 0.13 | -0.54 | 0.10 | 0.00 | 0.00 |
| Negative symptoms | -0.04 | 0.01 | -0.05 | 0.03 | 0.00 | 0.00 | 0.00 | 0.00 | 0.00 | 0.00 | -0.01 | 0.02 | -0.01 | 0.02 | 0.00 | 0.00 | -0.36 | 0.05 | 0.33 | 0.18 | 0.00 | 0.00 | -0.01 | 0.03 |
| Positive symptoms | 0.02 | 0.01 | -0.06 | 0.03 | 0.00 | 0.00 | 0.00 | 0.00 | 1.00 | 0.00 | 0.99 | 0.03 | -0.73 | 0.02 | -0.58 | 0.04 | -0.62 | 0.04 | -0.50 | 0.07 | 0.00 | 0.00 | 0.00 | 0.00 |
| Self-endangerment | -0.02 | 0.01 | 0.00 | 0.00 | 0.00 | 0.00 | 0.00 | 0.00 | 0.00 | 0.00 | 0.01 | 0.03 | -0.01 | 0.02 | 0.00 | 0.00 | 0.07 | 0.03 | 0.00 | 0.00 | 0.01 | 0.04 | 0.00 | 0.00 |
| Self-harm | -0.26 | 0.01 | -0.34 | 0.20 | 0.00 | 0.00 | 0.00 | 0.00 | 0.00 | 0.00 | 0.00 | 0.01 | -0.08 | 0.06 | 0.01 | 0.02 | 0.08 | 0.06 | 0.02 | 0.05 | 0.60 | 0.06 | 0.55 | 0.11 |
| Sleep | -0.04 | 0.02 | -0.04 | 0.03 | 0.00 | 0.00 | 0.00 | 0.00 | 0.00 | 0.00 | -0.01 | 0.04 | 0.03 | 0.05 | 0.01 | 0.02 | 0.10 | 0.07 | 0.03 | 0.06 | 0.00 | 0.00 | 0.00 | 0.00 |
| Suicidality | -0.28 | 0.01 | -0.03 | 0.03 | 0.00 | 0.00 | 0.00 | 0.00 | 0.00 | 0.00 | -0.01 | 0.04 | 0.17 | 0.12 | 0.25 | 0.04 | 0.11 | 0.04 | 0.03 | 0.05 | 0.58 | 0.01 | 0.02 | 0.04 |

Performance metrics across all diagnostic group-vs-other classifiers with 95% CIs. Feature weights given with standard error. Green columns are results from “best” prompts and red from “worst”. P-values for two-sided t-test comparison between “best” and “worst” models with α = 0.05. SUD - Substance Use Disorders.
