## Supplemental figures S4-S6 for "clickBrick Prompt Engineering: Optimizing Large Language Model Performance in Clinical Psychiatry"

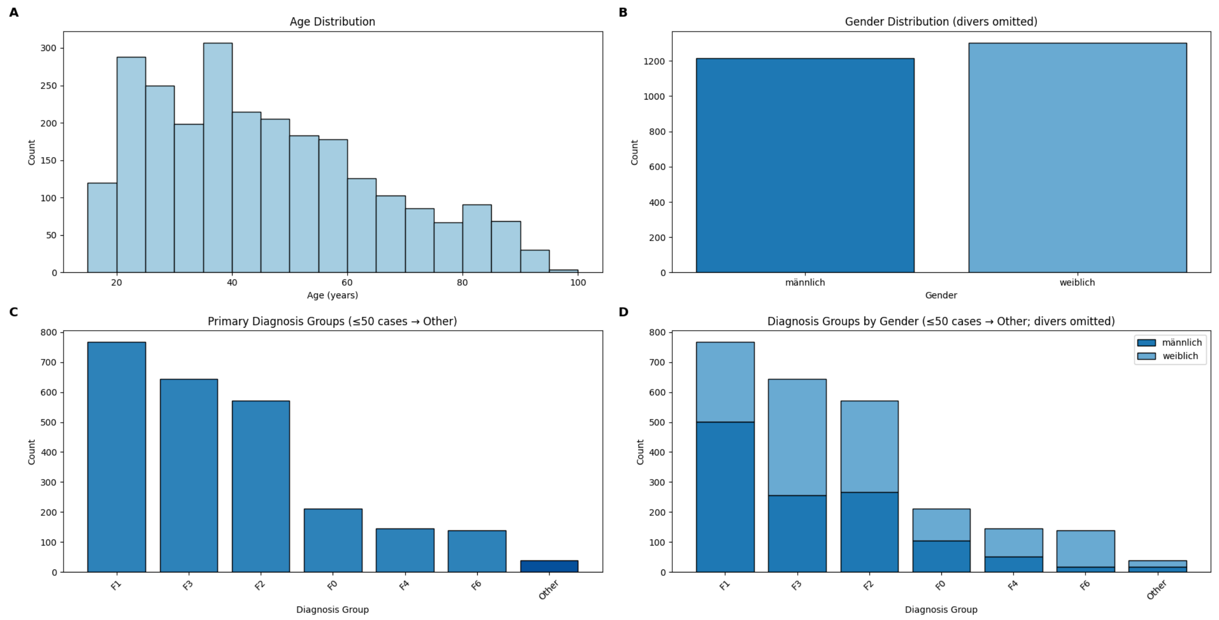


**Supplemental Figure S4**: Distribution of age, gender and primary diagnoses in 2.520 case samples from 1,692 patients. (A) Age distribution, mean age 45.2 ± 0.76 years, mean female age 45.95 ± 1.11 years, mean male age 44.43 ± 1.02 years. (B) Gender distribution, female: 1303 cases, male: 1215 cases, non-binary: 2 cases (not shown). (C) Primary diagnostic ICD-10 group, by frequency: F1 (substance use, any kind) 767 cases, F3 (affective disorders) 645 cases, F2 (Schizophrenia, schizotypal and delusional disorders) 572 cases, F0 (organic mental disorders, including dementia) 211 cases, F4 (anxiety disorders) 146 cases, F6 (disorders of adult personality and behavior) 141 cases, 38 other cases. (D) gender balance by main diagnostic groups (ratio = male/female, rounded): $F0_{ratio}$ = 1:1 (0.99), $F1_{ratio}$ = 2:1 (1.89), $F2_{ratio}$ = 1:1 (0.88), $F3_{ratio}$ = 2:3 (0.66), $F4_{ratio}$ = 1:2 (0.54), $F6_{ratio}$ = 1:7 (0.14).

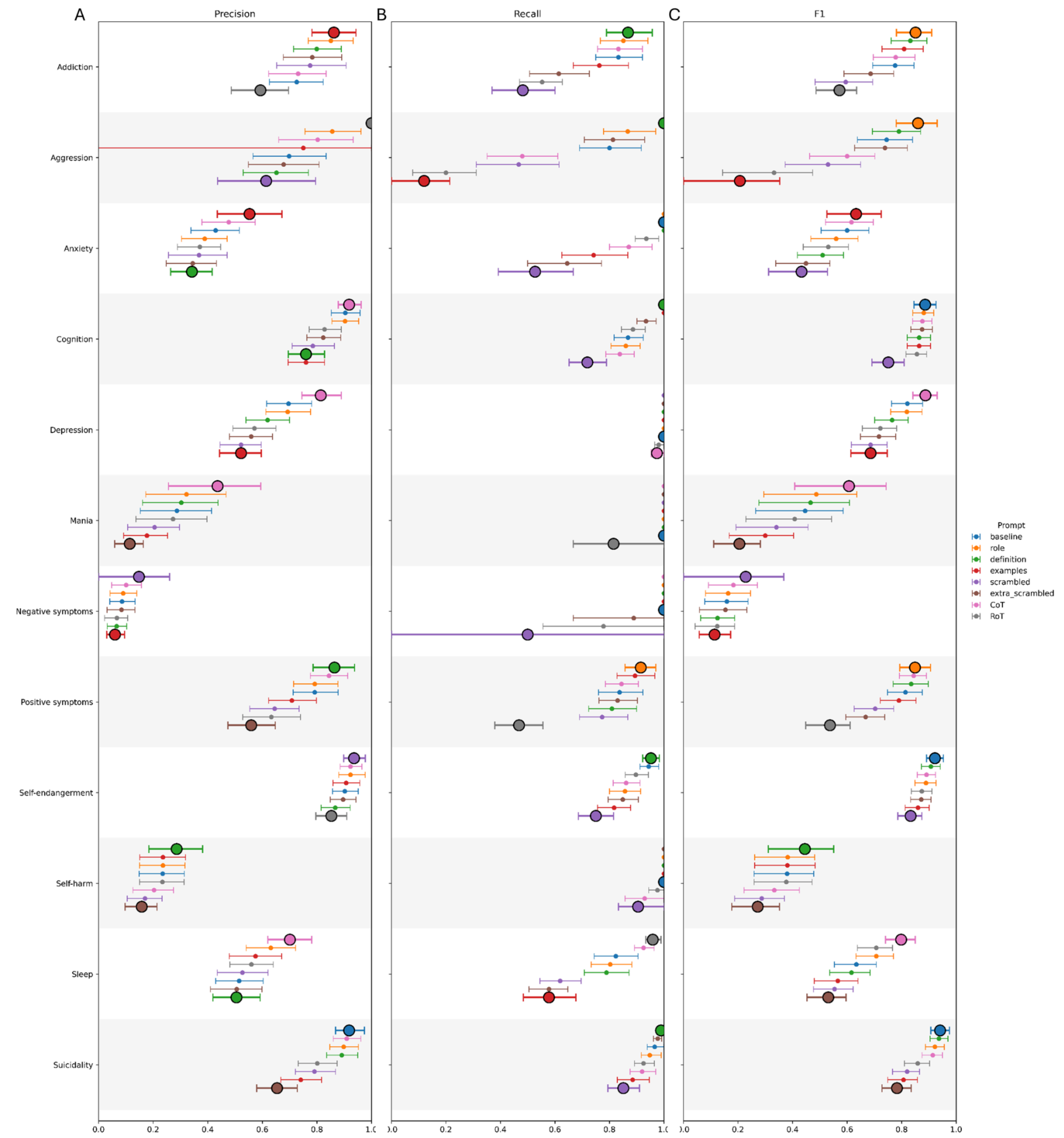


**Supplemental figure S5**: Performance metrics for all domains showing results for all prompts, highlighting best and worst prompting strategies (bold), error bars marking 95%-CIs from 2,000-fold bootstrapping across three independent experimental runs. (A) Precision, (B) Recall, (C) F1-score.

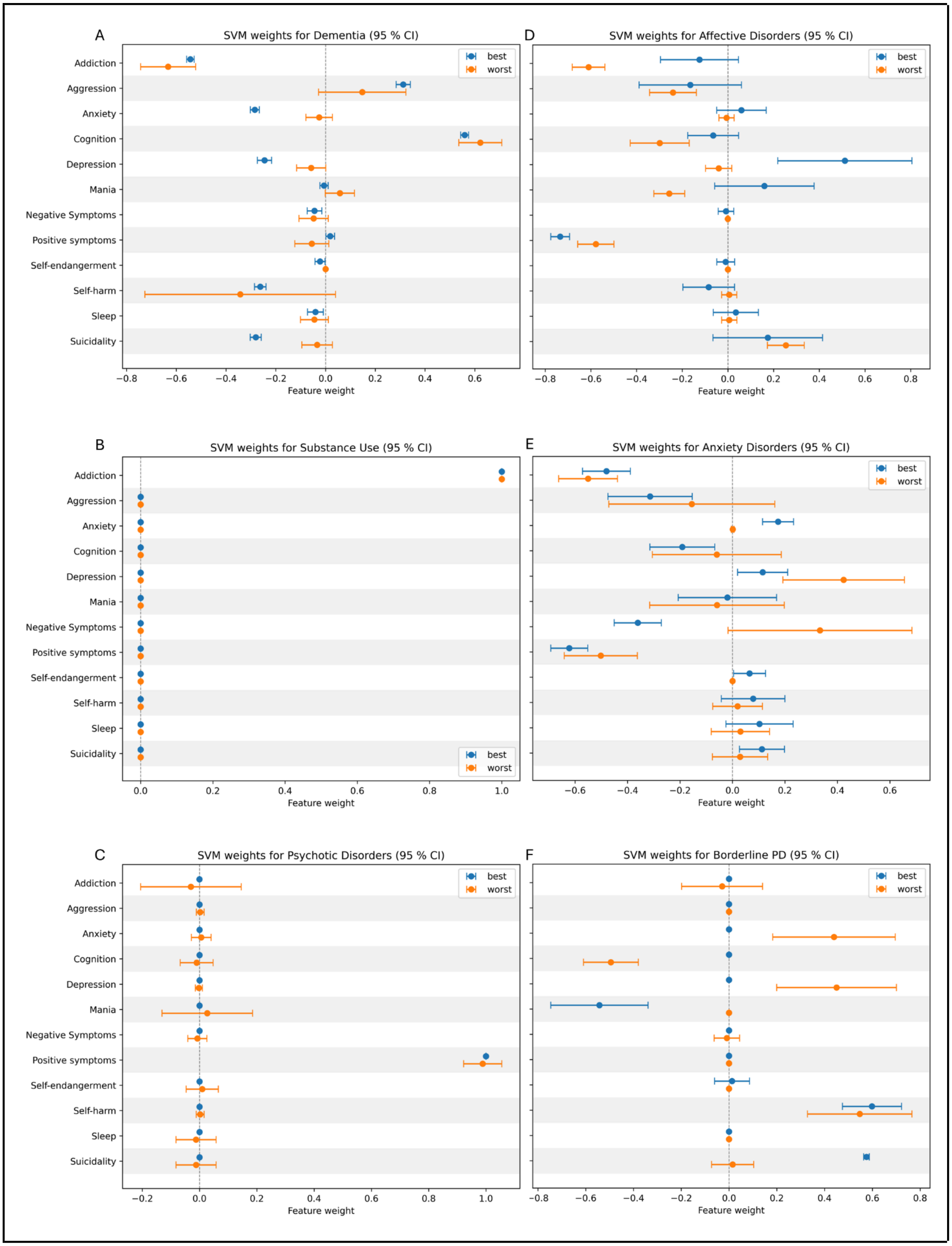
A
SVM weights for Dementia (95 % CI)
D
SVM weights for Affective Disorders (95 % CI)
B
SVM weights for Substance Use (95 % CI)
E
SVM weights for Anxiety Disorders (95 % CI)
C
SVM weights for Psychotic Disorders (95 % CI)
F
SVM weights for Borderline PD (95 % CI)
Addiction
Aggression
Anxiety
Cognition
Depression
Mania
Negative Symptoms
Positive symptoms
Self-endangerment
Self-harm
Sleep
Suicidality
best
worst
Feature weight

**Supplemental figure S6**: All feature weights for the machine learning classifiers. Shown for "best" (blue) vs. "worst" LLM-extraction strategies. Psychotic disorders and affective disorders as shown in figure 2B&C. Larger feature weight total values indicate stronger importance in SVM classification. Bars with whiskers indicate 95%-CIs. Corresponding BAcc as reference for (A) best: 82·25%, worst: 75·07%, (B) best: 78·86%, worst: 80·23%, (C) best: 74·97%, worst: 64·31%, (D) best: 69·67% worst: 70·85%, (E) best: 68·05% worst: 65·34%, (F) best: 83·11% worst: 75·84%.
